# A multicomponent behavior change intervention to promote walking in adults after traumatic brain injury: A Pilot Randomized Control Trial

**DOI:** 10.64898/2026.02.26.26347181

**Authors:** Timothy P. Morris, Emma M. Tinney, Skye Toral, Amanda O’Brien, Elda Gobena, Lexie Hackman, Mark C. Nwakamma, Madeleine Perko, Erin Orchard, Hannah Odom, Colette Chen, Jeremy Hwang, Noa Yedidia, Alexandra Stillman, Arthur F Kramer, Goretti España-Irla

## Abstract

**Background:** Sedentary behavior is highly prevalent following traumatic brain injury (TBI) and the cognitive and behavioral sequelae of TBI create barriers to adopting physical activity. Effective behavior change interventions in community-dwelling TBI survivors remain scarce.

**Purpose:** This study assessed the feasibility and compliance of a multicomponent intervention on walking behavior in adults with TBI and provides hypothesis-generating descriptives of the effect of this intervention vs an attention-matched usual-care inspired control on walking outcomes.

**Methods:** Fifty-six community-dwelling adults aged 40–80 years with a mild-to-moderate TBI diagnosed between 3 months and 15 years prior participated in a 12-week, two-arm pilot randomized control trial. Participants were randomized to either a planning, reminders, and micro-incentives intervention or a health advice control condition. Intervention participants completed weekly phone calls to plan five 30-minute walks for the following week, received daily text messages or email reminders on planned walk days, and earned small monetary incentives upon walk completion. Control participants received weekly health education calls. Feasibility was operationalized as recruitment, retention, and adverse event rates. Compliance was assessed as valid wear time of the activity tracker and adherence was operationalized as weekly completed phone calls. We provide mean, standard deviation and point estimates with 95% confidence intervals of outcome data (weekly walk counts, walking duration, and step counts; baseline and pre-to-post change in perceived health, motivation and self-efficacy for exercise) between groups to aid in future intervention development and generate hypotheses.

**Results:** Forty-eight participants completed the study (retention rate: 84.2%), with high phone call compliance in both groups (intervention: 98.4%; control: 98.1%) and zero serious adverse events. The intervention group walked more times (interaction aIRR = 1.12 per week, 95% CI [1.04, 1.20]), for longer (b = 8.00 mins per week, 95% CI [4.63, 12.97]) and took more steps (b = 514 steps per week, 95% CI [249, 779]) than the control group over the course of the intervention.

**Conclusions:** Remotely delivered, multicomponent walking interventions are possible candidate programs to fill a gap in community-based healthcare for individuals living with lifelong consequences of TBI. These data provide support for a larger, fully powered trial prior to implementation in as a post-rehabilitation program.

## Introduction

Once re-integrated back into the community, individuals who have sustained a traumatic brain injury (TBI) are susceptible to physical inactivity, placing them at greater long-term risk for cardiovascular disease, neurodegenerative, and affective disorders[1–4]. A critical gap in access to community-based healthcare programs can leave survivors of TBI without meaningful support to build and maintain healthy habits once reintegrated back into their communities.

Physical activity (PA) and exercise have been shown to be effective lifestyle strategies to improve cardiovascular health, cognitive and brain health[5], and favorable disease outcomes[6], as well as overall morbidity and mortality[7–9]. However, engaging in PA requires substantial cognitive resources to plan and make decisions about when, where, and how to be physically active[10]. This cognitive load becomes challenging when competing priorities in one’s environment demand attention, which is particularly difficult with mental fatigue following TBI[11]. Moreover, one needs to actively interrupt the automaticity of habitual sedentary behavior to initiate a pre-planned behavior, requiring conscious moment-to-moment awareness, as well as self-regulatory control[12]. Since the health benefits of PA are often delayed rather than immediately rewarding, individuals must rely on higher-order cognitive processes to overcome the natural inclination toward effort minimization (while maximizing rewards)[13,14]. This reward deficit creates an additional cognitive burden, as an individual must actively convince themself to engage in effortful behavior despite an absence of immediate, tangible benefits.

Multicomponent interventions to scaffold complex PA behaviors may be particularly effective in individuals with TBI. For example, planning can reduce the cognitive burden of behavior initiation by creating automatic if-then behavioral scripts[15]. Behavioral reminders (e.g., push notifications or text messages) can interrupt habitual sedentary behaviors and trigger pre-planned activities[16]. Small but timely incentives may provide proximal rewards that compete with the immediate rewards experienced from sedentary behavior[17]. Around ∼70% of PA research with TBI and stroke patients have tested the associations between PA and health outcomes, whereas only 6% have tested the effectiveness of interventions to promote independent PA behaviors, according to one review[18]. Indeed, the only systematic review of PA behavior change interventions in TBI report that the overall efficacy of specific self-management interventions remains inconclusive[18]. As such, evidence of interventions to promote PA adoption in TBI survivors is needed.

Here, we conducted a 12-week, two-arm, pilot randomized control trial of a virtually delivered intervention consisting of planning, reminders, and micro-incentives to promote walking in adults with TBI. The objectives of this pilot trial were to assess the feasibility, compliance, and preliminary efficacy of this intervention compared to a control group receiving only health advice. In an exploratory aim, we examined potential modifiers of walking behaviors in adults with TBI and whether increased walking habits contribute to improved health outcomes.

## Methods

Details of all study protocols are described below. This study received ethical approval from the local Institutional Review Board (#23-06-18). This manuscript was prepared under the CONSORT 2010 statement: extension to randomized pilot and feasibility trials[19].

### Study Design

This was a hybrid remote and in-person 12-week, two-arm pilot randomized control trial. Participants were invited to participate in a virtual or in-person baseline session followed by a remote intervention (Figure 1). After the baseline session, participants randomized (using block randomization of 4-6 per block) into either an active or control intervention by one of two study staff members (LH, ET). Randomization allocation was concealed using REDCap and participants were blinded to the study hypotheses but investigators were not blinded to allocation. A series of cognitive function and psychosocial questionnaires were collected at baseline (before randomization) and after the 12-week intervention, either in-person (if the participant was local) or over a HIPAA compliant zoom video call. Local participants were invited to participate in an imaging session including magnetic resonance imaging (MRI) and electroencephalography (EEG); however, these data are not presented here due to the small n (n=10 for MRI, n=14 for EEG). Outcome measures of PA were collected continuously over 12 weeks using a provided Fitbit Inspire 3.

**Figure 1.**
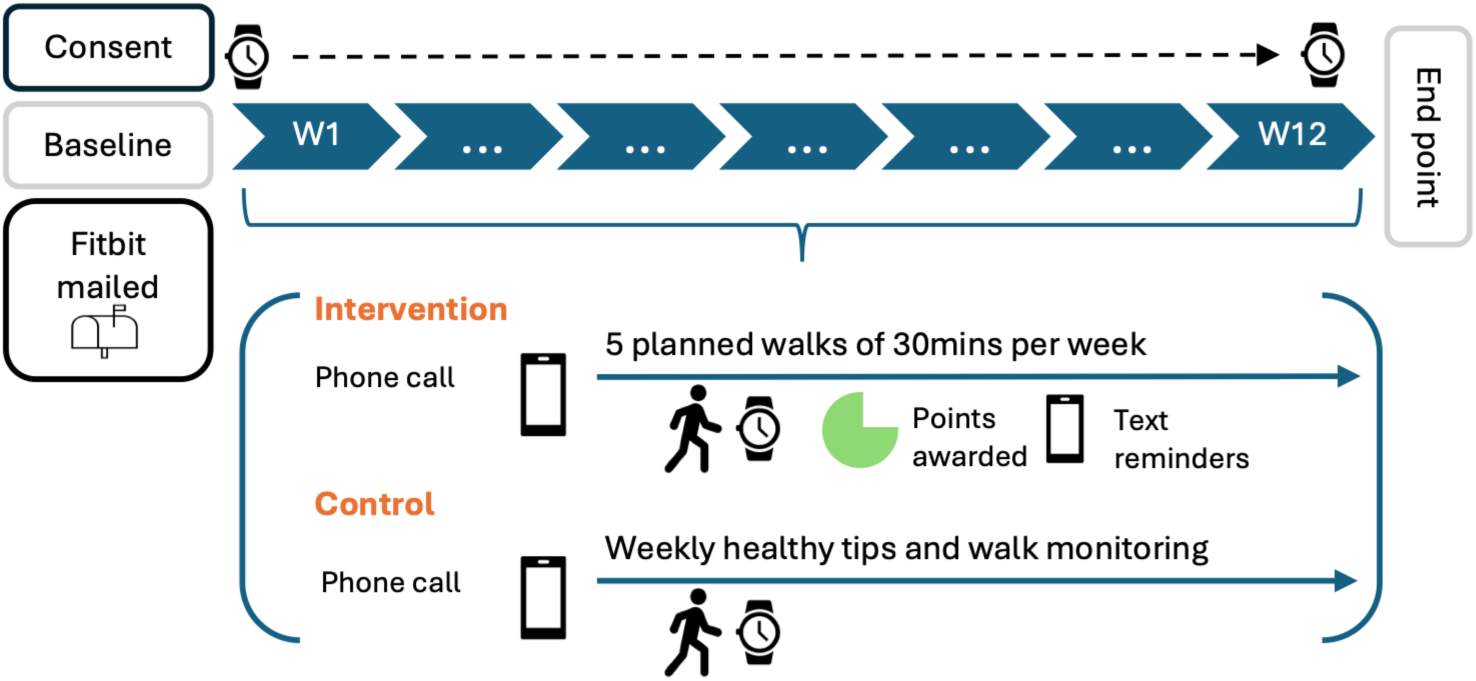
Study design.

### Participants

Adults aged 40-80 years with a diagnosis of mild or moderate TBI between 3 months and 15 years ago were recruited via local and national digital advertisements. An initial time since injury criteria of 2 to 10 years post injury was changed after consultation with TBI survivor groups to encompass more individuals living with long-term consequences of TBI. Participants were required to receive a formal mild or moderate TBI diagnosis by a physician (AS) using OSU-TBI-ID questionnaire [20], requiring a direct blow or impact to the head with either loss of consciousness of 0-30 minutes (mild) or >30 minutes and <24 hours (moderate) or post-traumatic amnesia of 0-1 day (mild) or >1 day and <7days (moderate). Additional inclusion criteria included the ability to walk without an assistive device. Exclusion criteria included a history of severe TBI (loss of consciousness >24 hours, post-traumatic amnesia > 7 days of a Glasgow Coma Scale of <9), current speech or physical therapy, mild cognitive impairment (as assessed by a mini-mental status exam score of ≤30) or neurological, neurodegenerative or cardiovascular disease.

### Sample size justification

The intended sample size for this pilot study was n=60 and derived pragmatically based on the feasibility of recruiting initially within a 1-year time frame. As a pilot study, we expected that n=60 would provide precise estimates of feasibility (drop-out probability, recruitment rate, intervention tolerability), and to quantify the precision and variance of the mean effect, which aids in the planning of larger, sufficiently powered efficacy trial[21,22]. We anticipated a 20% attrition rate and expected to have a final sample of 48, 24 in each group.

### Intervention

Planning, Reminders and Micro-incentives Intervention: During weekly phone calls with a study staff member participants randomized to this arm planned five 30-minute walks for the following seven days. Participants received a daily text message on planned walk days at 8am with a reminder of their daily plan and incentive notifications. Points were awarded when the activity tracker, connected to our propriety platform, confirmed walk completion, typically within 20 minutes to 1 hour (dependent upon the Fitbit API syncing). 500 points were awarded for each walk, with 1500 points converting to $1 at the end of the study. Participants were encouraged (during weekly phone calls) to track their progress on the study website (sFigure 1), view completed walks and point totals. Points were only awarded on planned walk days.

### Control Intervention

Those randomized to the control arm received similar weekly phone calls with information focusing on health education. This information was delivered with weekly phone calls from study staff members who delivered the control intervention only. Controls were provided with information in the form of “health tips” about the benefits of physical activity and other healthy behaviors. For example: “Walking 10,000 steps daily benefits cardiovascular health” (see Supplementary material 1 for completed list of weekly health advice). Participants were reminded to wear their Fitbit consistently and check the study platform for weekly health tips. Consistent scripts and staff training aimed to uniformly convey the content of the control conditions (Supplementary material 1) and match participant-staff contact time with that of the intervention group.

### Primary outcome measures

Feasibility was operationalized as recruitment, retention, and adverse event rates. Compliance was assessed via weekly phone call completion rates and adherence via Fitbit wear time. Efficacy outcomes included weekly walk counts (planned and total = planned and unplanned walks), walking duration, and step counts within walks. For analysis, a walk was counted if it met a qualifying criterion matching the prescribed 30-minute walks: a single walk of ≥30 minutes or two walks of 15–29.9 minutes on the same day (counted as one walk). Weekly walking duration and steps were summed over qualifying walks only. This criterion is stricter than the ≥27-minute threshold used to award points during the intervention (supplementary material 1). Detailed information on behavioral monitoring is found in supplementary material 1.

### Exploratory outcome variables

Exploratory outcomes were cognitive, motivation and self-efficacy scales collected at baseline and after the 12-week intervention over Zoom. Cognitive measures included perceived cognitive abilities (PROMIS Cognitive Function – Abilities Subset v2.0, Short Form 8a), verbal fluency (total words across four letter and category trials) and the Hopkins Verbal Learning Test (total recall, delayed recall, retention and delayed recognition discrimination index). Motivation was assessed with the six subscales of the Behavioral Regulation in Exercise Questionnaire 3 (BREQ-3)[23,24], which were also combined into a relative autonomy index (RAI; weighted sum of amotivation −3, external −2, introjected −1, identified +1, integrated +2 and intrinsic +3; range −24 to 24), and exercise self-efficacy with the Self-Efficacy for Exercise Questionnaire[25]. Detailed methodology for each variable is found in supplementary material 1.

### Statistical analyses

All analyses were performed in R 4.5.2 and included participants who completed the study (intervention *n* = 23, control *n* = 25). Demographics, feasibility and compliance outcomes are summarized descriptively, and call length is compared between groups as a rank-biserial correlation with 95% CI. For efficacy, weekly counts of qualifying walks were analyzed with Poisson generalized linear mixed models (log link), one for total walks and one comparing planned walks in the intervention group with total walks in controls, and weekly walking duration and step count with linear mixed-effects models (REML). All models included group, week (continuous, 1–12), their interaction, and covariates for age, sex, years of education, and standardized time since injury, fit in lme4 with a random intercept and random week slope for walk counts and a random intercept only for duration and steps. Participant-weeks outside 1.5 × the interquartile range of weekly duration or steps were excluded from those models. For one participant, time since injury was not recorded and was imputed at the median of the remaining participants (887 days). For one intervention participant every week exceeded the interquartile-range threshold and was removed from efficacy models. Walk-count effects are reported as adjusted incidence rate ratios and continuous outcomes as unstandardized coefficients, each with 95% Wald CIs; group contrasts at weeks 1, 6, and 12 were derived from estimated marginal means. No inferential hypothesis tests were performed, and no p-values are presented.

Three sets of exploratory models examined the 14 cognitive, motivation and self-efficacy scales each fit separately per scale. First, multilevel logistic regressions predicted whether a participant completed at least one qualifying walk on a given day (46 participants, 3,750 participant-days) from the z-scored baseline scale, adjusted for age, sex, years of education, time since injury, group and study month (1–3), with a random intercept per participant; effects are odds ratios per 1 SD with 95% Wald CIs. Second, pre-post change was modelled with linear mixed models (group × time, random intercept per participant; 48 participants), with both timepoints standardized on the baseline mean and SD; within-group changes and the group × time interaction are reported in baseline SD units. Given the exploratory aim, only 95% CIs are reported, without multiple-comparison correction.

## Results

### Feasibility

Between April 2024 and September 2025, 341 inquiries were received, 118 individuals passed initial screening and were prescreened for eligibility. Fifty-six participants (Table 1) were consented and randomized into either the intervention (*n*=28) or control arm (*n*=28). Eight participants (14.3%) dropped out of the study, five from the intervention group and three from the control group (Figure 2). Recruitment was timed to end in September 2025 relative to the grant end date (May 31^st^ 2026) resulting in 56 total enrollments. No serious adverse events related to study procedures were recorded throughout the study.

**Figure 2.**
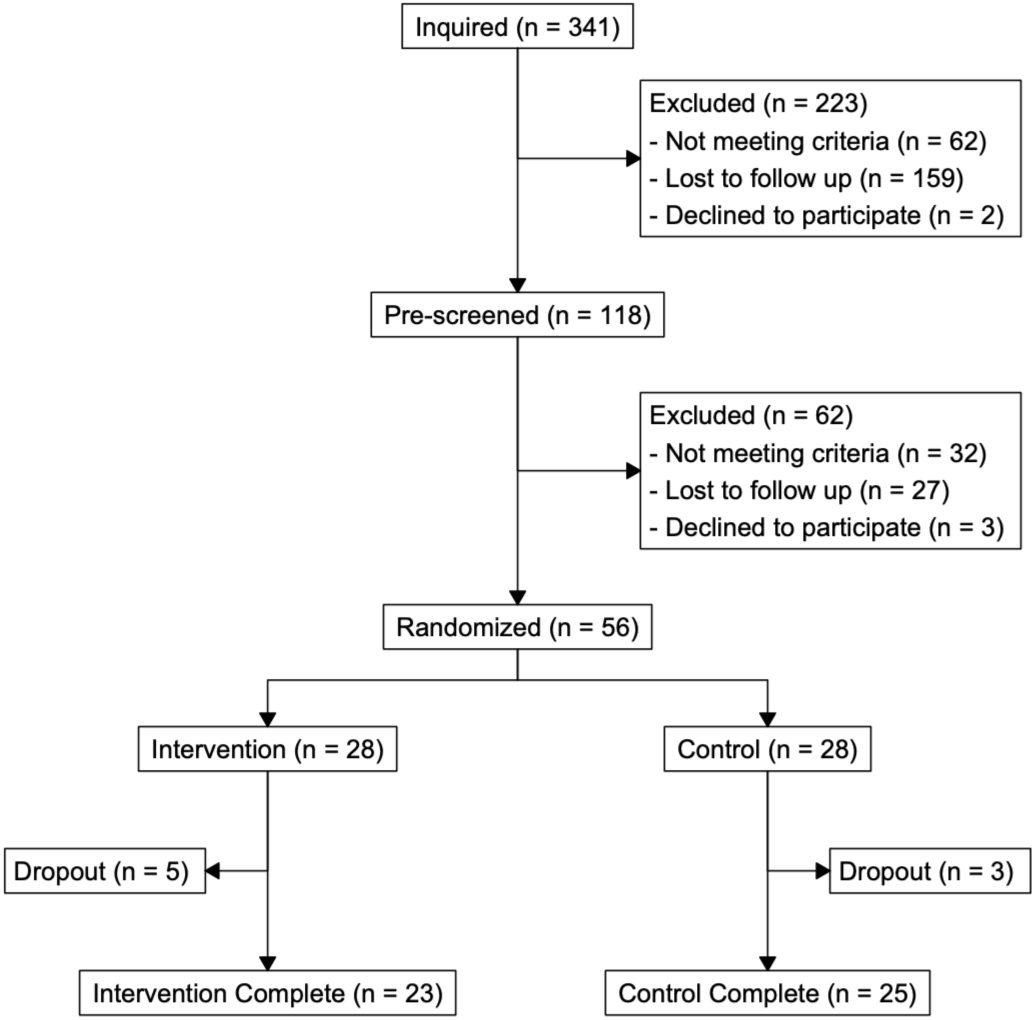
Consort diagram of participant enrollment, screening, randomization and retention.

**Table 1.** Baseline characteristics by group.

| **Characteristic** |  | **Overall** | **Intervention** | **Control** | **Dropout** |
|---|---|---|---|---|---|
| **n** |  | 56 | 23 | 25 | 8 |
| **Age (years) (mean (SD))** |  | 52.8 (8.9) | 53.0 (9.3) | 52.7 (8.5) | 52.6 (10.1) |
| **Biological Sex (%)** |  |  |  |  |  |
|  | Female | 48 (85.7) | 18 (78.3) | 23 (92.0) | 7 (87.5) |
|  | Male | 8 (14.3) | 5 (21.7) | 2 (8.0) | 1 (12.5) |
| **Gender Identity (%)** |  |  |  |  |  |
|  | Man | 8 (14.3) | 5 (21.7) | 2 (8.0) | 1 (12.5) |
|  | Woman | 48 (85.7) | 18 (78.3) | 23 (92.0) | 7 (87.5) |
| **Race (%)** |  |  |  |  |  |
|  | Asian | 1 (1.8) | 1 (4.3) | 0 (0.0) | 0 (0.0) |
|  | Black or African American | 3 (5.4) | 2 (8.7) | 1 (4.0) | 0 (0.0) |
|  | More Than One Race | 1 (1.8) | 1 (4.3) | 0 (0.0) | 0 (0.0) |
|  | Unknown | 1 (1.8) | 0 (0.0) | 1 (4.0) | 0 (0.0) |
|  | White | 50 (89.3) | 19 (82.6) | 23 (92.0) | 8 (100.0) |
| **Ethnicity (%)** |  |  |  |  |  |
|  | Hispanic or Latino | 2 (3.6) | 0 (0.0) | 1 (4.0) | 1 (12.5) |
|  | NOT Hispanic or Latino | 54 (96.4) | 23 (100.0) | 24 (96.0) | 7 (87.5) |
| **Time Since Injury (days) (mean (SD))** |  | 1246.5 (976.2) | 1219.7 (1099.2) | 1241.7 (896.5) | 1351.0 (943.7) |
| **Loss of Consciousness (%)** |  |  |  |  |  |
|  | No | 35 (62.5) | 14 (60.9) | 16 (64.0) | 5 (62.5) |
|  | Yes | 21 (37.5) | 9 (39.1) | 9 (36.0) | 3 (37.5) |
| **Memory Gap (%)** |  |  |  |  |  |
|  | No | 31 (55.4) | 12 (52.2) | 13 (52.0) | 6 (75.0) |
|  | Yes | 25 (44.6) | 11 (47.8) | 12 (48.0) | 2 (25.0) |

### Compliance

High weekly phone call compliance was found in both intervention (98.4%) and control (98.1%) groups. Phone call length was similar between groups (median [IQR], intervention: 4.3 [3.0, 6.2] minutes; control: 3.6 [3.1, 5.9] minutes; rank-biserial *r* = -0.04, 95% CI [-0.36, 0.28]; sTable 1 and sFigure 2A).

### Adherence

Participants wore their devices for on average 88.6% of days and no major differences in wear time was seen between groups (Table 2 and supplementary material 2).

**Table 2.** Activity tracker wear time compliance.

| **group** | **n** | **Days with Data Mean (SD)** | **% of Expected Days** | **Mean Daily Wear Time (hours)** | **Valid Days (>10hrs) Mean % (SD)** | **Daytime Wear % Mean (SD)** | **Nighttime Wear % Mean (SD)** |
|---|---|---|---|---|---|---|---|
| Control | 25 | 75.3 (22.4) | 87.5% | 18.9 (5.1) | 81.7 (25.5) | 67.4 (8.3) | 32.6 (8.3) |
| Intervention | 23 | 75.5 (11.7) | 89.7% | 16.4 (6.4) | 68.6 (30.1) | 67.1 (8.9) | 32.9 (8.9) |
| Overall | 48 | 75.4 (17.9) | 88.6% | 17.7 (5.8) | 75.4 (28.3) | 67.3 (8.5) | 32.7 (8.5) |

### Efficacy

The intervention showed an immediate effect, where at week 1, those in the intervention group completed more walks than controls (adjusted incidence rate ratio (aIRR) = 5.19, 95% CI [2.32, 11.6]) (Figure 3A&B). The control group (teal line) showed a declining trend, with predicted walks decreasing from 0.72 at week 1 to 0.26 at week 12 (aIRR per week = 0.911, 95% CI [0.853, 0.972]). In contrast, the intervention group maintained relatively stable walk counts across the study period. For total walks (orange line), intervention participants’ estimated walks remained between 3.76 (95% CI [2.08, 6.79]) at week 1 and 4.52 (95% CI [2.17, 9.42]) at week 12 (∼17.5x more than control). Planned walks (Figure 3A, red line) accounted for 60% of total intervention walks. The difference between intervention and control walks grew over the study period (interaction aIRR = 1.12 per week, 95% CI [1.04, 1.20], 89% power achieved using Wald-Based *posthoc* approximation), where intervention participants completed 5.2x more walks at week 1 and 17.5x more walks at week 12 (estimated marginal means are found in sTable 2). A comparison of planned walks and estimated marginal means for all models is found in supplementary material 2 and sTables 2,3.

**Figure 3.**
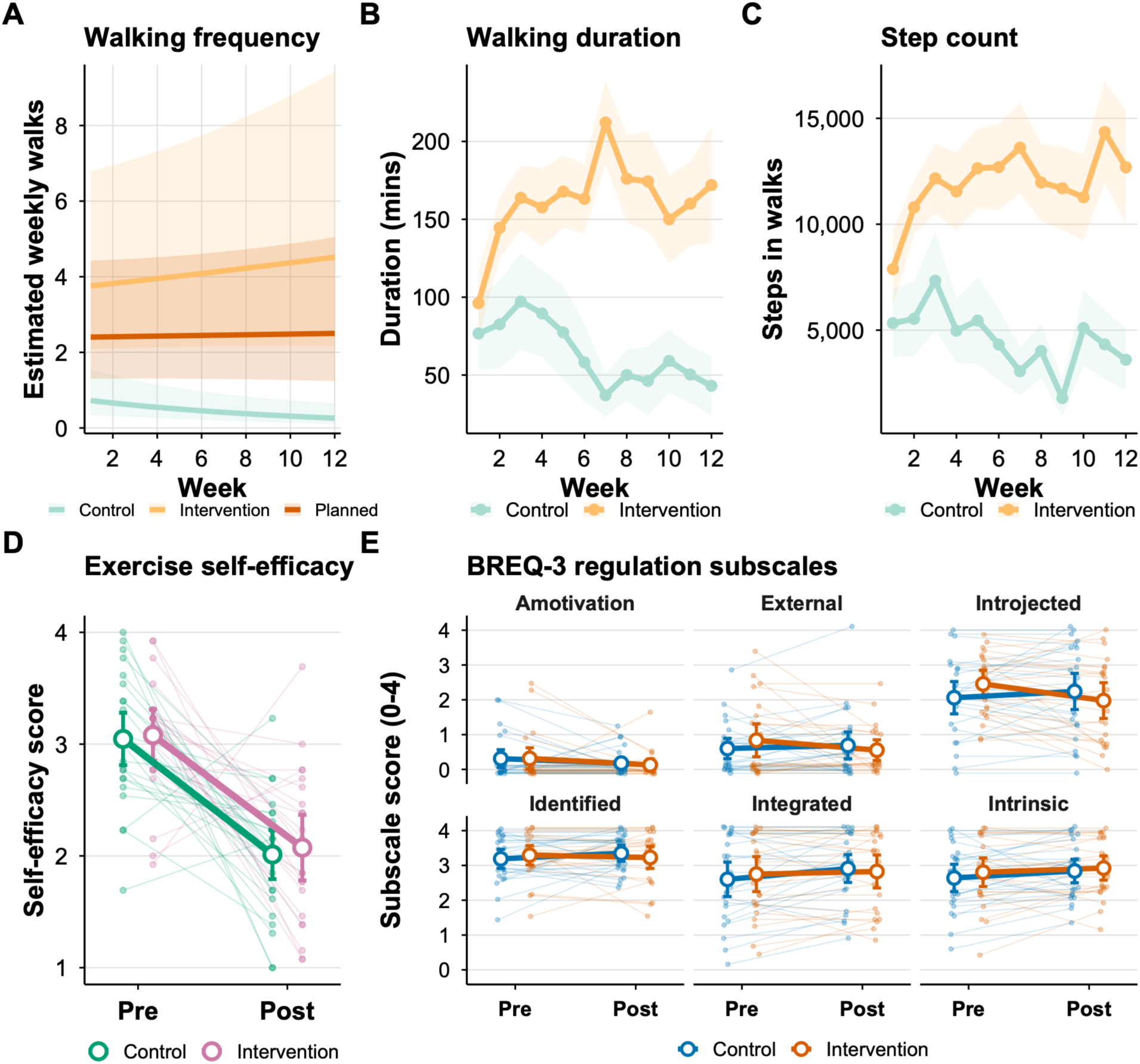
Multipaneled plot of intervention efficacy. (A) Model-estimated weekly walks with 95% CIs for control total walks, intervention total walks and the intervention’s planned-walk subset. (B) Observed mean weekly walking duration. (C) Observed mean weekly step count during walks. (D) Exercise self-efficacy from baseline to endpoint. (E) The six BREQ-3 regulation subscales from baseline to endpoint. Bands in panel A are model-based 95% CIs; bands in panels B and C are +/- 1 SE of the observed weekly means.

#### Duration

The intervention group walked on average 57.7 minutes more than the control group in the first intervention week (95% CI [1.3, 114.1]) (Figure 3C). The control group declined in walking duration over time (b = −4.58 minutes per week, 95% CI [−7.30, −1.85]). For every additional week, the intervention group walked an average of 8.80 minutes more than the control group during walk activities (b = 8.80, 95% CI [4.63, 12.97]). Estimated marginal means (sTable 4) show that during the final week, intervention participants (184.73 minutes) walked for an average 6.1x longer than control participants (30.24 minutes).

#### Steps

The intervention group on average took 4,093 more steps than the control group during walks at week 1 (95% CI [−311, 8,497]) (Figure 3D). A negative effect of week was observed in the control group, indicating a decline in step count over time (b = −250 steps per week, 95% CI [−423, −77]). For every additional week, the intervention group accumulated an average of 514 more steps during walks compared to the control group (b = 514, 95% CI [249, 779]). Estimated marginal means (sTable 5) shows that at week 12, intervention participants took 3.3x more steps during walks than the control group.

### Exploratory analyses – walking modifiers and pre-post changes

Higher baseline autonomous motivation was associated with greater odds of completing a qualifying walk (one walk of ≥30 minutes, or two walks of ≥15 minutes) on any given day, including intrinsic motivation (OR = 2.59 per SD, 95% CI [1.66, 4.04]), integrated regulation (OR = 1.87, 95% CI [1.15, 3.03]) and identified regulation (OR = 1.78, 95% CI [1.10, 2.87]), as was the BREQ-3 relative autonomy index (OR = 2.17, 95% CI [1.36, 3.46]). Amotivation, external regulation, introjected regulation and exercise self-efficacy were not clearly associated with walking (95% CIs including 1). Higher baseline perceived cognitive abilities (PROMIS Cognitive Function and HVLT delayed recall) were associated with lower odds of walking (OR = 0.51, 95% CI [0.31, 0.86]), whereas 95% CIs for objective cognitive measures (verbal fluency and HVLT) included 1 (sTable 7 and sFigure 3A).

From baseline to post-intervention, introjected regulation decreased in the intervention group relative to controls (group × time interaction = −0.63 baseline SD, 95% CI [−1.07, −0.20]; intervention: −0.46, 95% CI [−0.77, −0.15]; control: 0.17, 95% CI [−0.13, 0.47]). Interactions for the other BREQ-3 subscales and the relative autonomy index included zero. Exercise self-efficacy declined substantially in both groups (intervention: −1.84 SD, 95% CI [−2.47, −1.21]; control: −1.89 SD, 95% CI [−2.50, −1.29]; interaction = 0.05, 95% CI [−0.81, 0.91]). Group × time interactions for perceived cognitive abilities and all objective cognitive measures included zero (sTable 8 sFigure 3B).

## Discussion

In this 12-week, two-arm pilot randomized control trial, we report the feasibility, compliance and effect estimates of a remotely delivered planning, reminders and micro-incentives intervention to increase walking in community-dwelling adults with chronic mild-moderate TBI. We recruited participants across the continental United States, with a 15% attrition rate and high compliance throughout the weekly intervention and activity tracking. Effect estimates provide support for transitioning to a larger fully powered trial. However interesting signal within the motivation and self-efficacy scales provide some important insights into the design of any future trial.

This remote, technology-based intervention was well tolerated by individuals with a diagnosis of mild or moderate TBI. We report an attrition rate similar to health behavior change intervention studies[26] as well as high compliance with staff weekly phone calls and valid activity tracking wear time. Despite intervention participants wearing their device for fewer hours per day and having fewer valid wear days than controls, they still demonstrated substantially greater walking activity, suggesting that intervention participants were removing their devices after performing bouts of physical activity. From a practical and clinical standpoint, the intervention was delivered entirely remotely to participants distributed across the continental United States, without requiring in-person attendance. This is particularly relevant given that TBI survivors face numerous barriers to PA participation, including fatigue, transportation difficulties, limited access to adapted community programs, and lack of professional support[27,28]. Remote delivery circumvents some of these barriers, and combined with the low cost of the micro-incentives, suggests this intervention could offer a scalable and accessible program to promote physical activity once individuals have left in-person rehabilitation.

Physical activity may beget more physical activity through increased self-efficacy as repeated activity engagement builds confidence in capabilities[29–32], through increased autonomous motivation, as pleasurable experiences could shift externally motivated behavior to something that is intrinsically valued[33], or via improved cognitive function as chronic aerobic exercise itself can increase neurogenesis, neurotrophic factors and functional connectivity of networks that support cognitive performance[34,35]. However, whether those with a TBI retain the capacity for improvement in these constructs to a level sufficient to withdraw behavioral support is unknown. Moreover, whether multicomponent scaffolding interventions build these self-regulatory capabilities or create dependencies upon external scaffolding for behavior management is unclear. The preliminary hypothesis-generating outcomes analysis in this present study provides some important insights for future trial development. Exercise self-efficacy declined in both groups over the 12 weeks, suggesting that participants’ baseline perceptions of their exercise capability exceeded their actual capability upon walking regularly. This is a common in exercise-naïve older adults, but this also suggests that the intervention components themselves did nothing to increase self-efficacy for exercise in these participants. Future studies may consider phased introduction of the planned activities, with a goal of progressing to 5 weekly walks to increase perceptions of ones’ ability to carry out their planned activity while reducing disappointment and sense of failure when all 5 activities are not achieved. Of the motivation scales, amotivation, external regulation, introjected regulation and identified regulation decreased in the intervention group, and integrated regulation and intrinsic motivation increased. Together, these data provide some signal suggesting that participants in this study retain the capacity for improvement in exercise self-regulation and the intervention components themselves did not increase a drive to exercise only because of external rewards or internal pressures. Future trials may consider increasing the joy or satisfaction gained from the physical activity behavior in addition to the scaffolding components, perhaps through self-selection of the activity or through group-based social physical activities. Increasing integrated regulation and intrinsic motivation may improve the likelihood of continued physical activity engagement upon withdrawal of the intervention.

No pre-determined thresholds were set for progression to a larger fully powered trial. However, across all measures taken, we conclude that progression to a lager trial is warranted. We report acceptable recruitment and strong retention rates, missing data across outcome measures was minimal or zero, we saw strong compliance and adherence, and the protocol was delivered as intended, zero serious adverse events were recorded, and adverse event monitoring appeared adequate. Adequate variance in the primary outcome measure was also seen. Several lessons learnt however will improve the design and delivery of a larger trial. First, over half of all initial inquiries received were lost to follow-up. Ensuring improved contact with potential participants will improve recruitment rates and time to reach larger sample sizes, including improved representation of broad demographic groups. Second, the efficacy and outcome data suggest several amendments to the intervention design, including self-selection of physical activity to increase motivation and positive affect of the activities themselves as well as progressive increase in planned activities to minimize sense of failure early on in the program.

Several limitations to this study should be considered when interpreting the results. First, this was a pilot study and therefore not powered to test overall efficacy of the intervention. Effect sizes are reported with 95% confidence intervals and standard errors to inform the planning of a larger trial, and the study achieved only 72% power to detect the achieved effect size of the difference in walking bouts over time between groups (primary efficacy analysis). Second, participants varied considerably in time since injury, though this heterogeneity likely reflects the true diversity of the community-dwelling TBI population rather than a methodological limitation. Third, as a 12-week pilot trial, the long-term maintenance of the observed walking increases remains to be established, and future research should examine whether effects are sustained beyond the active intervention period. Fourth, the sample was not balanced by sex, with a predominance of female participants, and lacked racial and ethnic diversity, which limits the generalizability of the findings to more diverse TBI populations. Fifth, while Fitbit provided objective and continuous measurement of walking behavior, we only captured activities explicitly logged as walks, and incidental ambulatory activity or other activities (i.e. cycling or rowing) not recorded as a discrete walk event may have been missed. Finally, the intervention protocol has several modifiable features that may have constrained its efficacy but can be built upon in future larger trials, including the absence of detailed implementation intention planning (where, when and how), the use of generalized and fixed-time reminder messages, and uniform incentive structures applied across all participants regardless of individual reward sensitivity.

## Conclusions

This trial demonstrates that a remotely delivered, multicomponent behavior change intervention combining planning, reminders, and monetary micro-incentives is feasible, well-tolerated, and associated with increases in walking activity over 12 weeks in community dwelling adults with TBI. A larger, fully powered efficacy trial is warranted.

## Data Availability

All data produced in the present study are available upon reasonable request to the authors and will be made freely available in an online repository within 12 months of the final completion date of the study.

## Acknowledgments

This work was completed in part using the Discovery Cluster, supported by Northeastern University’s Research Computing team. We would like to thank all participants for their participation in this study, Dr. Therese O-Neil-Pirozzi for her help in recruiting participants as well as members of the Brain Injury Association of Massachusetts and the Brain Injury Association of America in promoting our study.

## Clinical Trials Registration

The study was registered on clincialtrials.gov before enrollment of the first participant (NCT06222502).

## Author contributions

TPM: Conceptualization, Data curation, Formal analysis, Funding acquisition, Investigation, Methodology, Project administration, Resources, Supervision, Writing—original draft. EMT: Conceptualization, Data curation, Investigation, Methodology, Project administration, Supervision, Visualization, Writing— review and editing. ST: Methodology, Formal analysis, Investigation. AOB: Data curation, Investigation, Writing—reviewing and editing. EB: Data curation, Investigation, Writing—reviewing and editing. LH: Data curation, Project administration, Supervision. MCN: Data curation, Investigation, Methodology, Supervision, Visualization, Writing— reviewing and editing. MP: Data curation, Investigation, Writing—reviewing and editing. EO: Data curation, Investigation, Writing—reviewing and editing. HO: Data curation, Investigation, Writing—reviewing and editing. HO: Data curation, Investigation, Writing—reviewing and editing. CC: Data curation, Investigation, Writing—reviewing and editing. JH: Data curation, Investigation, Writing—reviewing and editing. NY: Data curation, Investigation, Writing—reviewing and editing. AMS: Conceptualization, Investigation, Writing—reviewing and editing. AFK: Conceptualization, Methodology, Writing—review and editing. GEI: Conceptualization, Methodology, Data curation, Formal analysis, Supervision, Writing—reviewing and editing.

## Transparency statements

The study was pre-registered at clinicaltrials.gov: NCT06222502. The analysis plan was not formally pre-registered. De-identified data from this study will be available on Zenodo within 12 months of the primary study completion (May 31^st^ 2026). Analytic code used to conduct the analyses presented in this study are available in a public archive at https://github.com/cnelabneu/Morris26_walkingtbi.git. Some of the materials used to conduct the study are presented in a public archive at https://github.com/cnelabneu/roybal.git and others can be found in the supplementary materials.

