## supplementary material for "A multicomponent behavior change intervention to promote walking in adults after traumatic brain injury: A Pilot Randomized Control Trial"

### Supplementary Materials

#### Supplementary Material 1. Expanded methodology

Intervention Monitoring:

Two password-protected websites were used to deliver and monitor the intervention: 1) a staff website for inputting and modifying participant schedules, and pulling Fitbit data automatically in near-real time, and 2) a participant website displaying a walk calendar, point tallies, and color-coded completion status (supplementary figure 1). The system connected to Fitbit's Web API for near-real-time data collection and used Twilio to send automated reminders. Data typically synced within 20 minutes after a walk, points were awarded automatically upon walk completion, and individual data were downloaded for offline analysis. Participants were monitored for adverse events via weekly phone calls, with any health or safety concerns documented, reviewed by the principal investigator, and classified according to National Institute on Aging guidelines. An Inspire 3 Fitbit was mailed after baseline, with setup scheduled 7–14 days later via Zoom (~30 minutes). Study staff created a de-identified Google account for each participant prior to setup, which covered platform familiarization and device and application configuration. Daily data were extracted from participants' Fitbit Google Accounts, including minute-level heart rate, daily step counts, and walk activity logs (start time, duration, steps, distance, heart rate, caloric expenditure), filtered to each participant's active intervention period. Study weeks were defined as 7-day periods beginning from each participant's first weekly phone call, which was combined with the setup call. Device wear time was calculated using heart rate data as the primary indicator of wear, following Baroudi and colleagues[30], as the number of unique minutes with heart rate data divided by 1,440, multiplied by 100. Daytime (7:00AM–11:00PM) versus nighttime (11:00PM–7:00AM) wear time percentages were also calculated to characterize wearing patterns. Walk data were extracted from Fitbit activity logs (labeled "Walk" only). During the intervention, walks were validated for points if they comprised either a single walk ≥27 minutes or two walks of 15–27 minutes on the same day; the ≥27-minute threshold was chosen to reduce participant frustration for walks of 27–29.99 minutes. For analysis, a stricter criterion matching the prescribed 30-minute walks was applied in both groups: a qualifying walk was a single walk of ≥30 minutes or two walks of 15–29.9 minutes on the same day (counted as one walk), and weekly walking duration and steps were summed over qualifying walks only. For intervention participants, the first completed walk on a planned day was classified as "planned," with additional walks on that day or any unscheduled day classified as "unplanned," ensuring planned walk counts could not exceed scheduled walks while capturing activity beyond the protocol. For control participants, all completed walks contributed to their total walk count.

Control arm weekly script and health tips:

Control Participant
Hello [participant name].
How has your week been going so far?
How is the Fitbit? Have you been able to charge it so far? Have you had any issues or discomfort with it?
Great do you have any questions about Fitbit maintenance?

Thank you for letting me know. Are you ready to hear this weeks' health education tip? [health tip of the week].
Have a nice week; I look forward to hearing speaking with you again next week.

Strategies + Information Points:

1. You can make small and easy changes in your daily life to get more active! You can get off the bus or train one stop early and walk the rest of the way or even try parking at the far end of the parking lot.
a. Walking decreases the risk or severity of various health outcomes such as cardiovascular and cerebrovascular diseases, type 2 diabetes mellitus, cognitive impairment and dementia, while also improving mental well-being, sleep, and longevity.
2. Stay motivated and enhance the enjoyment of your walking activity by teaming up with a walking buddy. There are also lots of local walking or hiking groups to join that help you to stay active and meet new people.
a. Combining physical activities with social interaction has been linked to maintaining brain health. Several studies have suggested that social interaction can lead to a higher quality of life, greater satisfaction, and a lower risk of cognitive impairment and dementia over time.
3. Enhance relaxation and mental clarity by incorporating meditation into your walking routine, where focus is placed on your breathing and each step taken. There are also lots of available free guided meditations either through YouTube, Spotify, or other dedicated meditation apps!
a. Recent studies suggest that incorporating mindfulness practices, such as meditation, gratitude, and mindful eating and communication, into your daily routine may lead to reduced stress, improved mental health, and increased overall well-being.
4. Ensure you stay hydrated during your walks by bringing along a small water bottle, especially for longer periods of walking. The recommended amount for men is 3,000ml and 2,200ml for women per day. (For reference a normal plastic water bottle is about 500!)
a. Adults who stay well-hydrated appear to be healthier, develop fewer chronic conditions, such as heart and lung disease, and live longer.
5. Keep entertained during your walks by considering a fun or interesting podcast to listen to, offering a pleasant diversion along the way! There's topics ranging from animals to sports and everything in between!
a. Podcasts can boost mental health, reduce stress, and improve your active listening skills. They're also great for learning new things and reducing screen time.
6. Enjoy a change of scenery by visiting local nature parks or arboretums, where you can experience beautiful views while taking your walk.
a. Consciously spending time outside, especially in a wooded area, has been shown to improve mood, increase vitamin D levels, and improve the immune system's functioning.
7. Enhance the enjoyment of your walk by finding or creating a music playlist featuring a diverse selection of songs. You can get creative by incorporating your movie soundtracks or mixing your favorite genres!
a. Studies have shown that music increases motivation, distracts from feelings of fatigue and even improves the efficiency of workouts.
8. Instead of emailing or texting your neighbors or co-workers, walk to their office or house and speak with them when possible! This gives great face-to-face interaction

and helps you stay active during the day!
a. Routine physical activity, whether as part of a job or as a leisure activity, is associated with reductions in the incidence of specific cancers, in particular colon and breast cancer.
9. To keep your mind engaged, you can create a walking bingo game or mentally play the alphabet game by finding something along your route that begins with A, B, C and so on! It'll not only keep your mind active, but help you pay more attention and appreciate your surroundings.
a. Walking boosts immune function. Walking can help protect you during cold and flu season. A study (found that people who walked regularly) had 43% fewer sick days than those who exercised once a week or less.
10. Treat yourself to something enjoyable, like a small indulgence or an episode of your favorite show, after each walk. It can give you something to look forward to if you're getting tired during the walk!
a. Physical activity stimulates many brain chemicals that may leave you feeling happier, more relaxed and less anxious.
11. Discover the excitement of exploring new and interesting locations by taking walks in unexplored parts of your town or city. For example, within Boston, you can take the Freedom Trail or a self-guided art walk.
a. Studies have shown that being in a new place could increase your empathy, energy, focus, and attention, which helps build strong memories.
12. If you typically like to talk to friends and family on the phone, try taking the phone calls while walking. It can give you something to keep your mind occupied while also making the time spent walking fly by.
a. It's been found that walking at least 1 mile a day significantly enhanced the volume of several brain areas, including the frontal lobe, which is involved with problem solving and reasoning... By walking at least 1 mile a day, they had reduced their risk of cognitive impairment by approximately 50%.

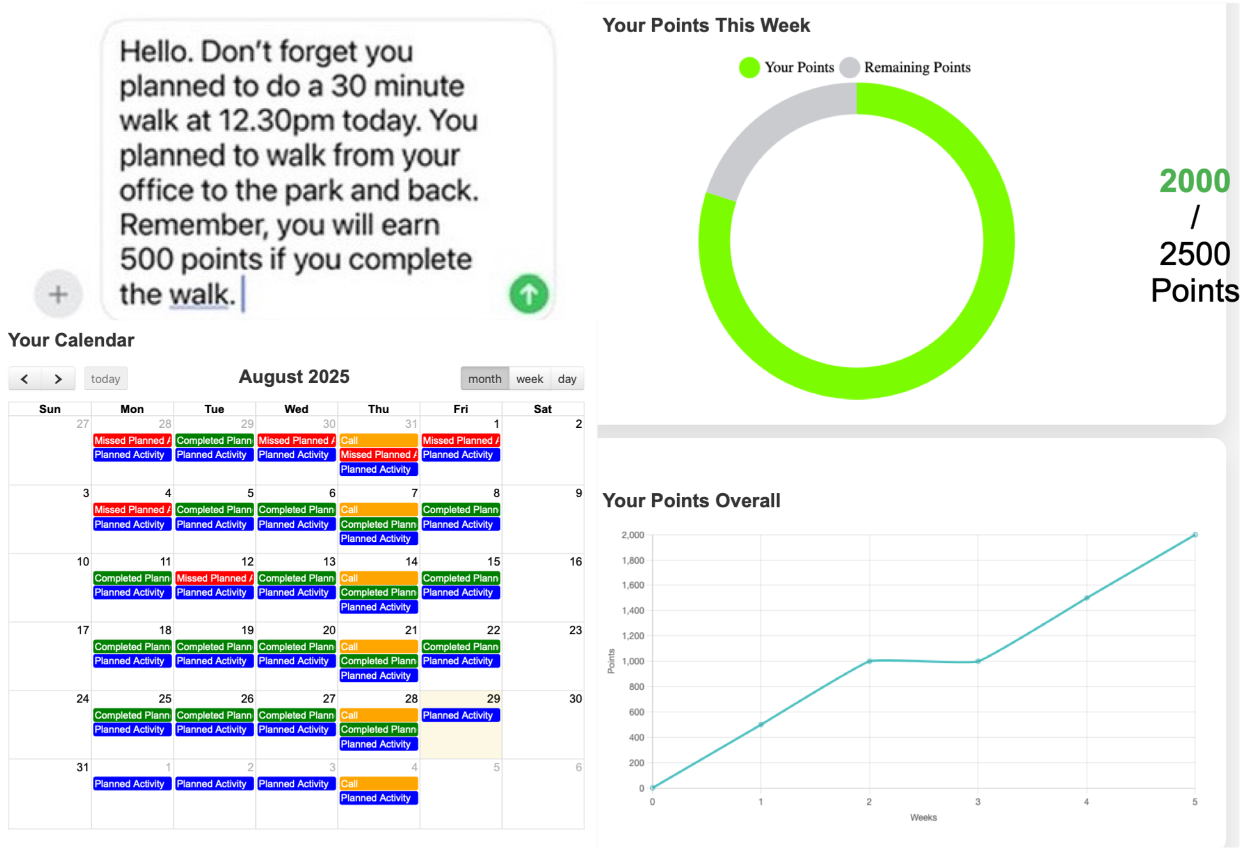


**sFigure 1**. Illustration of the participant-specific study platform. Participants received either a text message or email reminder on the day of planned walks (top left). They could access their online profile and see a calendar of their planned walks (in blue), when they had the next call scheduled (orange), and a past history of whether they completed or not each walk (green for completed, red for missed). Their points tally was depicted here also, with a weekly power bar filling up as points accumulated and a study progression plot of the accumulated points over time.

A series of cognitive and psychosocial questionnaires were administered at baseline and post-intervention. Some of these variables were not analyzed in this manuscript as they were out of scope of the primary aims. Others are presented to further future intervention development  All collected measures during the course of this study are detailed below for transparency.

Cognitive function:
Verbal Fluency[31] and Hopkins Verbal Learning Test[32] were administered at baseline and endpoint. Verbal fluency was scored as the total number of words across four trials (letters F, A and S and the category animals at baseline; letters C, F and L and the category fruits at endpoint). HVLT scores comprised total recall (sum of trials 1–3), delayed recall (trial 4), retention (trial 4 / trial 3 × 100) and the delayed recognition discrimination index, with an alternate form at endpoint. Because stimuli differed between timepoints, within-group changes in these measures are confounded with form difficulty. A paper version of the Trail Making Test was administered to participants who completed their baseline session in person but not presented here due to the small n (n=10). All cognitive testing was administered by two trained research staff (EMT and LH).

Psychosocial questionnaires:

Patient-Reported Outcomes Measurement Information System (PROMIS)[29–31] questionnaires were assessed at both baseline (before intervention assignment) and following the intervention. PROMIS instruments are developed and maintained by the National Institutes of Health (NIH) and use item response theory (IRT) to provide precise, standardized measurement of health-related quality of life across diverse populations. Perceived global health was assessed using the PROMIS Global Health Scale v1.2 (PROMIS-10), a 10-item self-report measure which evaluates physical, mental, and social health perceptions. The 10 items aggregate into two summary subscales: the Global Physical Health (GPH) subscale and the Global Mental Health (GMH) subscale. The GPH subscale is defined by items assessing physical health, physical function, pain, and fatigue. The GMH subscale is defined by items assessing quality of life, mental health, satisfaction with social activities, and emotional distress. Subjective cognitive functioning was assessed using the PROMIS Cognitive Function – Abilities Subset v2.0, Short Form 8a. This 8-item self-report measure evaluates patient-perceived cognitive abilities across domains including mental acuity, memory, processing speed, concentration, and the ability to track and manage daily tasks. The Abilities Subset uses exclusively positively-worded items that capture perceived cognitive competence. All scores are reported as T-scores calibrated to the U.S. general population (mean = 50, standard deviation = 10), and higher scores indicate better functioning for all three scales.

A health and demographic survey collected information on demographic variables and past and present medical history.

The Behavioral Regulation in Exercise Questionnaire-3 (BREQ-3), a 24-item self-report measure assessing the continuum of behavioral regulations for exercise motivation, grounded in Self-Determination Theory. Items are rated on a 5-point Likert scale (0 = "not true for me" to 4 = "very true for me") and grouped into six subscales: amotivation (4 items), external regulation (4 items), introjected regulation (4 items), identified regulation (4 items), integrated regulation (4 items), and intrinsic regulation (4 items). These subscales reflect a continuum from controlled to autonomous motivation, with higher autonomous regulation (identified, integrated, intrinsic) associated with more sustained exercise behavior. The BREQ-3 extended the original BREQ and BREQ-2 by adding the integrated regulation subscale, which reflects exercise behavior that is fully assimilated into one's values and sense of self.

A Self-Efficacy for Exercise Questionnaire, a self-report measure assessing participants' confidence in their ability to exercise regularly under various barriers (e.g., fatigue, bad weather, low motivation). Participants rate their confidence on a scale, with higher scores reflecting greater exercise self-efficacy.

The Pittsburgh Sleep Quality Index (PSQI) is PSQI is a validated self-report questionnaire yielding information about overall sleep quality, latency, duration, efficiency, disturbances, medication use, and the effects of daytime function across a previous 30-day interval. The total score ranges from 0 to 21, with higher scores indicating worse sleep quality.

The International Physical Activity Questionnaire (IPAQ), a self-report measure assessing physical activity across four domains (work, transport, domestic, and leisure) over the past seven days. Responses are used to calculate total weekly metabolic equivalent of task (MET) minutes, categorizing participants as low, moderate, or high activity.

The Mediterranean Diet Adherence Screener (MEDAS), a 14-item self-report screener assessing adherence to a Mediterranean dietary pattern, including consumption of olive oil, fish, legumes, fruits, vegetables, and red meat. Scores range from 0–14, with higher scores indicating greater adherence.

The Patient Health Questionnaire (PHQ-9), a 9-item self-report measure assessing the frequency of depressive symptoms over the past two weeks, based on DSM diagnostic criteria. Scores range from 0–27, with established cutoffs for mild, moderate, moderately severe, and severe depression.

The Generalized Anxiety Disorder Questionnaire (GAD-7), a 7-item self-report measure assessing the frequency of anxiety symptoms over the past two weeks. Scores range from 0–21, with cutoffs for mild, moderate, and severe anxiety, and a score of ≥10 commonly used as a clinically meaningful threshold.

#### Supplementary Material 2. Expanded results

Feasibility:

Five of these dropouts were for motivational reasons, two for musculoskeletal injuries not due to study procedures, and one for issues associated with a major hurricane. See the consort diagram in Figure 2 for details.

Seven adverse events not related to study procedures were recorded. Four participants reported musculoskeletal injuries sustained in the home, two reported head injuries also sustained in the home, and one reported a major illness.

Compliance:

Participants in the intervention group had a higher % of days, with data showing 89.7% in this group compared to 87.5% in the control group (Table 2). Participants in the control group on average wore their Fitbit device for longer on any given day and had more valid days with >10 hours of wear time. However, participants in both groups wore their device during the day (7.00am to 11.00pm) for ~70% of the total wear time. See supplementary material 5 and supplementary figure 2B for a comparison of mean daily wear time between group.

sTable 1. Phone call adherence and length. Completed and missed coaching calls against the 11 scheduled, and the length of completed calls. Call length is a per-participant mean, so the median and IQR are across participants rather than across calls.

| Group | N | Completed, mean (SD) | Completion rate | Missed, mean (SD) | Length (min), mean (SD) | Length (min), median (IQR) |
|---|---|---|---|---|---|---|
| **Control** | 25 | 10.6 (0.8) | 98.1% | 0.2 (0.4) | 4.7 (2.4) | 3.6 (3.1-5.9) |
| **Intervention** | 23 | 10.7 (0.5) | 98.4% | 0.2 (0.5) | 4.8 (2.2) | 4.3 (3.0-6.2) |
| **Overall** | 48 | 10.7 (0.7) | 98.2% | 0.2 (0.4) | 4.7 (2.3) | 4.0 (3.0-6.1) |

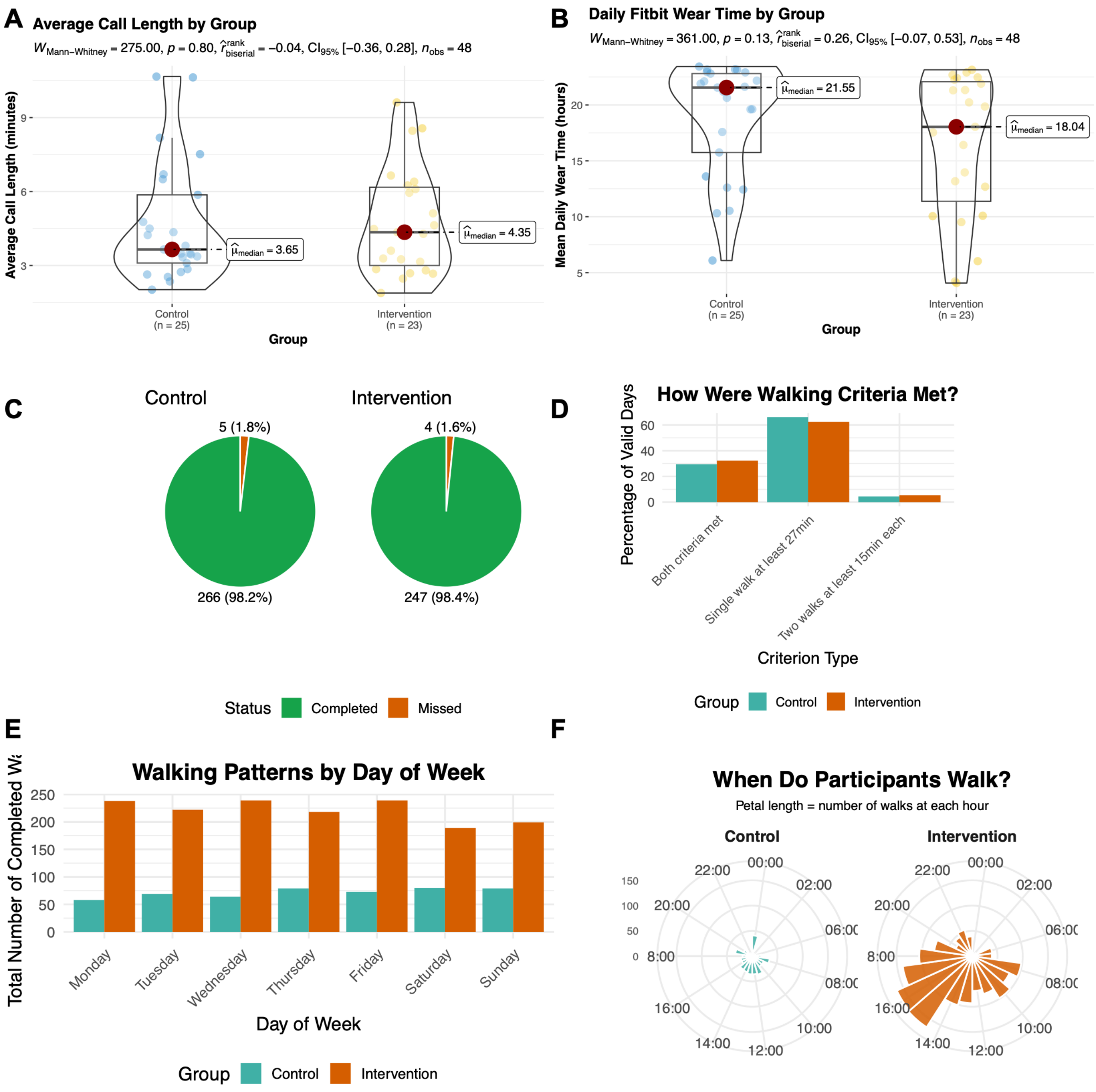


**sFigure 2**. Compliance and Exploration of Walking criteria. Panel A: A statistical comparison of average call length by group. Panel B: Shows the average daily wear time difference between groups. Panel C illustrates the total (green) and missed (red) weekly phone calls by group). Panel D represents the percent of days where a walk was either completed as a single walk of >27mins, two walks of 15+ mins or both (the participant walked for 27mins and then again in the afternoon walked twice for 15+ mins. Panel E shows the breakdown of walks by day of the week. Panel F illustrates the time of day when walks were completed, broken down by group.

Efficacy:

A numerical majority of completed walks were >27 minutes in length (Supplementary Figure 2D). Most walks happened between 2pm and 6pm in the evenings, but fewer total walks were completed on a given weekend (Supplementary Figure 2E and 2F).

sTable 2. Estimated marginal means of weekly walks (planned and unplanned) at reference weeks 1, 6 and 12. Model 1: Poisson GLMM, group x week, adjusted for age, sex, education and standardised days since injury, with random intercept and slope for week.

| Week | Control (95% CI) | Intervention (95% CI) | Rate ratio (95% CI) |
|---|---|---|---|
| 1 | 0.72 (0.34, 1.55) | 3.76 (2.08, 6.79) | 5.19 (2.32, 11.58) |
| 6 | 0.45 (0.21, 0.97) | 4.09 (2.16, 7.74) | 9.00 (3.87, 20.93) |
| 12 | 0.26 (0.10, 0.65) | 4.52 (2.17, 9.42) | 17.45 (6.12, 49.76) |

Assessment of planned walks only:

A Poisson generalized linear mixed model examined planned walk counts (intervention group only) compared to all walks in the control group over 12 weeks, adjusting for age, sex, education, and days since injury. Random intercepts and slopes for week were included to account for individual variability.

At week 1, the intervention group's planned walks were higher than the control group's total walks (adjusted incidence rate ratio [aIRR] = 3.95, 95% CI [1.75, 8.93]). Among control participants, walk counts declined over time (aIRR = 0.927, 95% CI [0.867, 0.991]), representing a 7.3% weekly decrease. The intervention-by-week interaction (aIRR = 1.08, 95% CI [1.01, 1.16]) indicated that the gap between planned intervention walks and control walks widened over time.

These findings suggest that even when considering only planned walks (excluding spontaneous walking), intervention participants maintained substantially higher walking activity than controls throughout the study period (estimated marginal means in sTable 3). Covariate estimates were imprecise for age (aIRR = 0.966, 95% CI [0.923, 1.01]), sex (aIRR = 1.38, 95% CI [0.427, 4.49]), education (aIRR = 1.01, 95% CI [0.846, 1.20]), and days since injury (aIRR = 1.09, 95% CI [0.737, 1.60]).

sTable 3. Estimated marginal means of planned walks (intervention) versus total walks (control). Model 2: same specification as Model 1, with the outcome set to planned walks in the intervention arm and all walks in the control arm.

| Week | Control, total (95% CI) | Intervention, planned (95% CI) | Rate ratio (95% CI) |
|---|---|---|---|
| 1 | 0.61 (0.28, 1.30) | 2.40 (1.30, 4.43) | 3.95 (1.75, 8.93) |
| 6 | 0.42 (0.19, 0.90) | 2.45 (1.30, 4.61) | 5.88 (2.59, 13.37) |
| 12 | 0.26 (0.10, 0.69) | 2.50 (1.24, 5.05) | 9.48 (3.51, 25.63) |

sTable 4. Estimated marginal means of weekly walk duration (minutes). Model 3: linear mixed model, random intercept only, Kenward-Roger degrees of freedom.

| Week | Control (95% CI) | Intervention (95% CI) | Difference in minutes (95% CI) |
|---|---|---|---|
| 1 | 80.6 (28.1, 133.0) | 138.2 (89.7, 186.8) | 57.7 (1.3, 114.1) |
| 6 | 57.7 (7.2, 108.2) | 159.4 (113.3, 205.5) | 101.7 (49.3, 154.1) |
| 12 | 30.2 (-22.4, 82.8) | 184.7 (135.1, 234.3) | 154.5 (97.1, 211.9) |

sTable 5. Estimated marginal means of weekly step count during walks. Model 4: linear mixed model, random intercept only, Kenward-Roger degrees of freedom.

| Week | Control (95% CI) | Intervention (95% CI) | Difference in steps (95% CI) |
|---|---|---|---|
| 1 | 6,952 (2,798, 11,105) | 11,045 (7,224, 14,865) | 4,093 (-311, 8,497) |
| 6 | 5,701 (1,646, 9,756) | 12,365 (8,667, 16,063) | 6,664 (2,464, 10,863) |
| 12 | 4,200 (39, 8,361) | 13,949 (10,068, 17,829) | 9,749 (5,291, 14,207) |

sTable 6. Fixed effects for all four walking models. Walk counts are adjusted incidence rate ratios; duration and step count are unstandardized coefficients. 95% Wald confidence intervals.

| Term | Estimate (95% CI) |
|---|---|
| **Model 1. Total walks - adjusted incidence rate ratio** | |
| Group (intervention) | 4.646 (2.063, 10.466) |
| Week | 0.911 (0.853, 0.972) |
| Age | 0.963 (0.922, 1.007) |
| Sex (male) | 1.852 (0.611, 5.615) |
| Years of education | 1.050 (0.879, 1.255) |
| Time since injury (SD) | 1.095 (0.754, 1.590) |
| Group x week | 1.117 (1.043, 1.195) |
| **Model 2. Planned vs total walks - adjusted incidence rate ratio** | |
| Group (intervention) | 3.648 (1.588, 8.382) |
| Week | 0.927 (0.867, 0.991) |
| Age | 0.966 (0.923, 1.010) |
| Sex (male) | 1.384 (0.427, 4.490) |
| Years of education | 1.010 (0.846, 1.205) |
| Time since injury (SD) | 1.085 (0.737, 1.597) |
| Group x week | 1.083 (1.012, 1.159) |
| **Model 3. Weekly walking duration, minutes - unstandardized b** | |
| Group (intervention) | 48.88 (-8.11, 105.87) |

sTable 6. Fixed effects for all four walking models. Walk counts are adjusted incidence rate ratios; duration and step count are unstandardized coefficients. 95% Wald confidence intervals.

| Term | Estimate (95% CI) |
|---|---|
| Week | -4.58 (-7.29, -1.86) |
| Age | -0.28 (-3.22, 2.65) |
| Sex (male) | -14.63 (-96.72, 67.47) |
| Years of education | -3.68 (-14.75, 7.39) |
| Time since injury (SD) | 20.04 (-5.74, 45.82) |
| Group x week | 8.80 (4.64, 12.96) |
| **Model 4. Weekly steps in walks - unstandardized b** | |
| Group (intervention) | 3579 (-812, 7970) |
| Week | -250 (-423, -78) |
| Age | 12 (-223, 248) |
| Sex (male) | 2202 (-4383, 8787) |
| Years of education | 117 (-771, 1004) |
| Time since injury (SD) | 547 (-1514, 2608) |
| Group x week | 514 (250, 779) |

#### *Exploratory analysis tables and figures:*

sTable 7. Baseline scales as predictors of daily walking. Odds ratios per 1 SD from separate multilevel logistic models (outcome: day with a qualifying walk of ≥30 minutes or two walks of ≥15 minutes), each adjusted for age, sex, education, days since injury, group and study month.

| Scale | OR | 95% CI |
|---|---|---|
| **Cognition** | | |
| PROMIS Cognitive Function | 0.51 | [0.31, 0.86] |
| Verbal fluency | 0.84 | [0.49, 1.45] |
| HVLT total recall | 0.68 | [0.41, 1.12] |
| HVLT delayed recall | 0.62 | [0.38, 1.00] |
| HVLT retention | 0.70 | [0.42, 1.16] |
| HVLT recognition discrimination | 0.95 | [0.57, 1.59] |
| **Motivation (BREQ-3)** | | |

sTable 7. Baseline scales as predictors of daily walking. Odds ratios per 1 SD from separate multilevel logistic models (outcome: day with a qualifying walk of ≥30 minutes or two walks of ≥15 minutes), each adjusted for age, sex, education, days since injury, group and study month.

| Scale | OR | 95% CI |
|---|---|---|
| **Amotivation** | 0.60 | [0.35, 1.00] |
| **External regulation** | 0.73 | [0.42, 1.25] |
| **Introjected regulation** | 1.27 | [0.75, 2.13] |
| **Identified regulation** | 1.78 | [1.10, 2.87] |
| **Integrated regulation** | 1.87 | [1.15, 3.03] |
| **Intrinsic motivation** | 2.59 | [1.66, 4.04] |
| **BREQ-3 composite** | | |
| **Relative autonomy index** | 2.17 | [1.36, 3.46] |
| **Self-efficacy** | | |
| **Exercise self-efficacy** | 0.94 | [0.56, 1.57] |

sTable 8. Pre-post change by group in baseline SD units (estimate [95% CI]). Within-group changes are simple slopes (endpoint minus baseline); the interaction is the difference between them (intervention minus control).

| Scale | Control change | Intervention change | Group × time interaction |
|---|---|---|---|
| **Cognition** | | | |
| **PROMIS Cognitive Function** | 0.20 [−0.23, 0.62] | 0.27 [−0.17, 0.72] | 0.08 [−0.54, 0.69] |
| **Verbal fluency** | −0.16 [−0.42, 0.09] | −0.41 [−0.68, −0.15] | −0.25 [−0.62, 0.12] |
| **HVLT total recall** | 0.27 [−0.15, 0.68] | 0.25 [−0.19, 0.68] | −0.02 [−0.62, 0.58] |
| **HVLT delayed recall** | 0.35 [−0.06, 0.76] | 0.17 [−0.27, 0.60] | −0.18 [−0.78, 0.41] |
| **HVLT retention** | 0.01 [−0.44, 0.46] | 0.03 [−0.44, 0.50] | 0.01 [−0.64, 0.67] |
| **HVLT recognition discrimination** | 0.10 [−0.37, 0.57] | 0.14 [−0.36, 0.64] | 0.04 [−0.64, 0.73] |
| **Motivation (BREQ-3)** | | | |
| **Amotivation** | −0.20 [−0.60, 0.21] | −0.28 [−0.70, 0.14] | −0.08 [−0.67, 0.50] |
| **External regulation** | 0.10 [−0.25, 0.45] | −0.31 [−0.67, 0.05] | −0.41 [−0.91, 0.10] |
| **Introjected regulation** | 0.17 [−0.13, 0.47] | −0.46 [−0.77, −0.15] | −0.63 [−1.07, −0.20] |

sTable 8. Pre-post change by group in baseline SD units (estimate [95% CI]). Within-group changes are simple slopes (endpoint minus baseline); the interaction is the difference between them (intervention minus control).

| Scale | Control change | Intervention change | Group × time interaction |
|---|---|---|---|
| **Identified regulation** | 0.25 [−0.14, 0.63] | −0.10 [−0.50, 0.30] | −0.35 [−0.91, 0.21] |
| **Integrated regulation** | 0.27 [0.04, 0.49] | 0.07 [−0.17, 0.30] | −0.20 [−0.53, 0.13] |
| **Intrinsic motivation** | 0.21 [−0.06, 0.48] | 0.13 [−0.16, 0.41] | −0.09 [−0.48, 0.31] |
| **BREQ-3 composite** | | | |
| **Relative autonomy index** | 0.19 [−0.07, 0.45] | 0.27 [0.003, 0.54] | 0.08 [−0.29, 0.46] |
| **Self-efficacy** | | | |
| **Exercise self-efficacy** | −1.89 [−2.50, −1.29] | −1.84 [−2.47, −1.21] | 0.05 [−0.81, 0.91] |

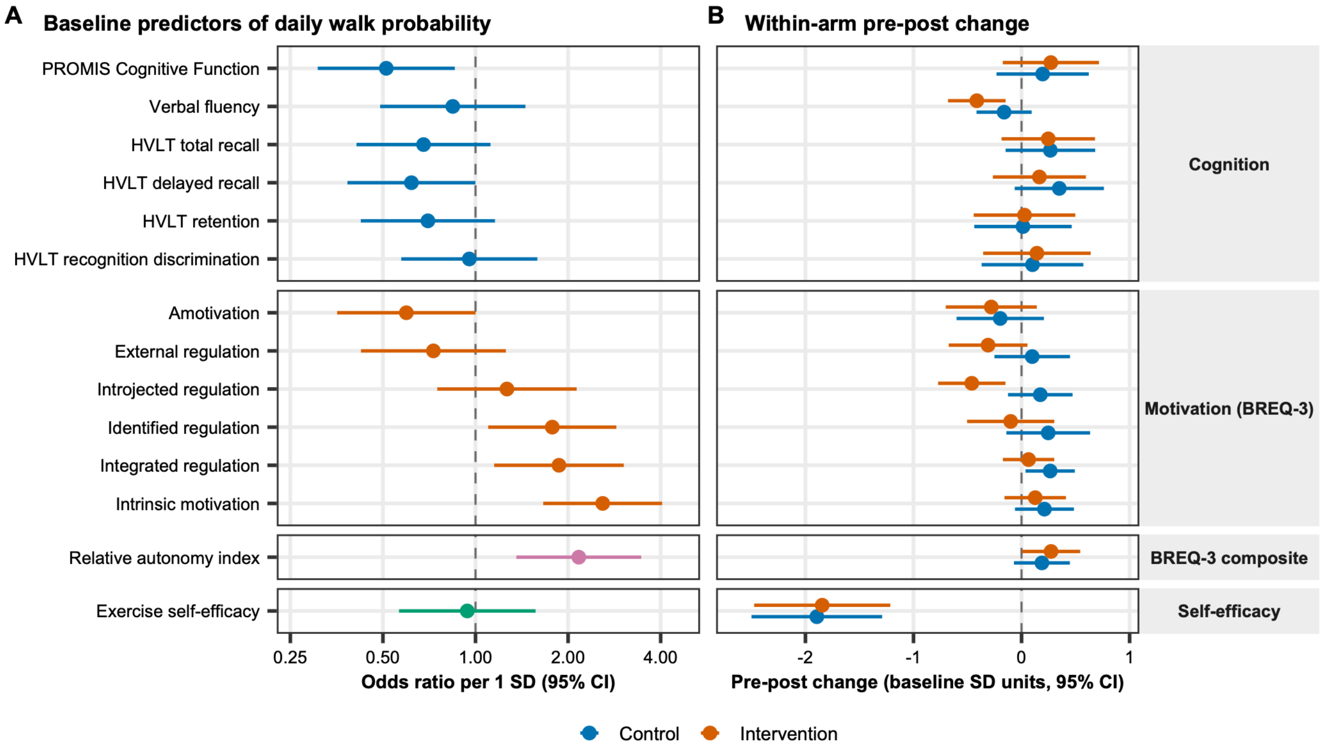


**sFigure 3.** Point estimate and 95% Cis for baseline predictors of walking (A) and the within arm pre-post change in each outcome measure (B).
